# Modelling the potential contributions of polygenic risk stratification to cost-effective screening for prostate cancer

**DOI:** 10.1101/2025.05.19.25327897

**Authors:** Padraig Dixon, Jonathan Aning, Richard M. Martin, Mark Clements

## Abstract

**Background:** Prostate cancer (PCa) is one of the most common cancers globally. The potential for polygenic risk scores (PRSs) to improve risk stratification for early detection of PCA in screening interventions is of considerable interest. We evaluated the cost-effectiveness of PRS-guided screening strategies.

**Methods:** The Prostata microsimulation model, calibrated to UK-specific epidemiological and clinical data, simulated individual life histories, including disease onset and progression, tumour characteristics by Gleason score, and metastatic status. The model incorporated both measured and unmeasured components of polygenic risk for incident PCa, and included ancestry-specific strata. Screening strategies were compared against a no-screening baseline. We modelled one-off, age-based prostate specific antigen screening at 50, 60, or 70 years, as well as repeated uniform screening every 4 or 2 years from age 50 to 69 with and without PRS stratification.

**Results:** Quality-adjusted life years were similar across all screening strategies, while differences in costs were more pronounced. A “no screening” strategy had the lowest lifetime cost of all strategies and (very marginally) the shortest life expectancy. Realistic PRS implementations were dominated (less effective and more expensive) in all scenarios, and may not provide greater cost-effectiveness than a single PSA screen at age 50 or compared to no screening at all.

**Conclusion:** Our study found little evidence that PRSs would be cost-effective in pragmatic PCa screening settings.

## 1 Introduction

Prostate cancer (PCa) is the second most common form of cancer in men globally, with almost 1.5 million incident cases recorded worldwide in 2022 (1). A key challenge in PCa screening is accurately identifying men with clinically significant disease while minimizing the overtreatment of indolent tumours (2–5).

Germline genetic variation influences the risk of incident PCa (6, 7), which suggests a possible role for polygenic risk scores (PRSs) in risk stratification (8–21). PRSs are a form of summary genetic data that reflect cumulative effect of numerous genetic variants associated with an individual’s genetic predisposition to disease. The variants that comprise a PRS are fixed at conception and can be assayed at any point in life (22, 23).

Cost-effectiveness is an essential requirement for any new tool such as PRS to be considered in population level screening programmes (24–26). To date, health economic research on PCa PRS in screening interventions has been sparse, and economic research on the potential for PRSs in screening has been recommended as a priority (11). A recent systematic review (20) identified clear evidence gaps in the contribution of PRSs in cost-effective cancer screening. The need for research accounting for differential availability and predictive capacity of PRSs by ancestry has been highlighted (27).

The objective of this paper was to assess the potential impact that the use of polygenic risk scores (PRSs) may have on the cost-effectiveness of population-wide PCa screening. To do so, we used a modified version of the Prostata PCa natural history microsimulation model (28). This model simulates life histories of individuals at risk of PCa and accounts for changes in PSA scores before and after disease onset, progression from localised to advanced PCa, and tumour stage (T-stage) and metastatic stage (M-stage) by Gleason score.

## 2 Methods

### 2.1 Microsimulation model

The version of the Prostata model we analyzed was calibrated by Keeney et al (29) to UK-specific PCa parameters including patterns of PSA testing, PCa incidence by age and Gleason score, as well as healthcare costs and quality of life parameters. The model’s mortality predictions were calibrated and validated against mortality rate ratios reported in ten-year follow up of the CAP trial (4) and 16-year follow-up from European Randomised Study of Screening for Prostate Cancer (30). (Treatment effects were based on data from the Prostate Testing for Cancer and Treatment (ProtecT) trial (31). A health economic analysis plan was not developed for this updated version of the model.

Polygenic susceptibility to PCa incidence was modelled as frailty, following a log-normal distribution to reflect individual relative risk within the population. The average measured polygenic risk was normalized to 1, with a mean of −0.68/2 and variance of 0.68 on the natural log scale. The model accounted for unmeasured polygenic risk, introducing variability in PCa risk. This unmeasured risk had a mean of −1.14/2 and a variance of 1.14 on the log scale, keeping the population’s average risk centred at 1. The distinction between measured and unmeasured frailty enable inferences about how increased knowledge of “true” but knowable polygenic risk could affect risk assessment.

We modelled a hypothetical cohort of 10 million men born in the UK in 1950 and followed their natural history of PCa over their projected lifetimes. This individual-based modelling approach allows us to evaluate the cost-effectiveness of different PCa screening strategies across the entire life course of the cohort. For instance, the model can simulate the introduction of screening at any particular age and track outcomes over a lifetime, rather than being limited to assessing impacts only in older age groups, such as men in their sixties or seventies.

To ensure comparability across different model runs, natural histories of specific simulated men were controlled by common random numbers. This means that simulated men will experience the same underlying randomness (e.g., same disease progression risk) except for what is directly affected by the different screening interventions. This ensures that any comparisons across strategies are always meaningfully “like for like”. For these common random numbers, we first sampled for the total (or “true”) frailty, then calculated the conditional log(PRS) mean and standard deviation, sampled the PRS frailty from the conditional PRS distribution, and then calculated the remaining (or “observed”) frailty from the total frailty divided by the PRS frailty.

To incorporate strata for ancestry, we assumed that a birth cohort in 1950 would include approximately 5% “Black” ancestry and 5% “Asian” ancestry with hazard ratios for cancer onset of 2.62 and 0.50, respectively. This mix of ancestries very roughly corresponds to ethnic distribution of the UK in the 2020s of men aged approximately 70 (given the assumed birth year of 1950). For example, 93% of adults aged 65-74 in the 2021 UK census reported White ethnicity, while 4% reported “Asian ancestry” amongst adults aged over 65 and 1.4% reporting “Black”. We have therefore over-represented the proportion of individuals in these minority ethnic groups in the model, compared to the 2021 population structure, but these ancestral groups will grow in size as younger individuals age and become eligible for the types of PCa screening interventions we model.

Accounting for these differences, we otherwise assumed a common PRS and a common screening risk threshold across ancestries.

### 2.2 Screening strategies compared and PRS scenarios modelled

We studied screening strategies as follows from an NHS/health service perspective as follows. We considered no screening as a comparator against which other strategies could be compared, and to reflect current UK practice, which does not recommend population-wide screening for PCa.

We then studied three strategies which considered a single one-off uniform screening strategy starting at ages 50, 60 and 70 respectively. These strategies are “uniform” in the sense that they apply to everyone meeting the single respective age criteria, and are one-off in the sense that they are not repeated for any reason. We also modelled uniform screening with repeated screening from age 50 to age 69 every 4 years and every 2 years respectively. In all these strategies, men receive a PSA test at the initial screen. If the PSA score is ≥3.0 ng/ml , they receive a multiparametric MRI scan and (if indicated) a transperineal biopsy.

We incorporated polygenic risk-informed stratified screening as follows. For each of these scenarios, we instead assumed an initial screening using a PRS. Men were assumed to start PSA testing if they were in the top 50% of PRS disease risk, after which they would then receive further investigations (if indicated) in the sequence of PSA test, MRI, and biopsy. For each of these “PRS first” strategies, we considered two scenarios for this polygenic risk stratified screening, which we refer to as “upper bound”, and “pragmatic”, as follows.

Scenario 1: “UPPER-BOUND” This anti-conservative scenario describes an unobtainable ideal for PRS-stratified screening. We assumed that a screening programme has access to a true PRS; that is, it assumed all germline genetic contributions (whether common or rare and including epistatic effects) to incident PCA risk have been discovered and are calculable for all individuals in a hypothetical screening programme. We assumed that the cost of obtaining and using the PRS is £0. Overall, this scenario offers an upper bound, “best chance” for PRS risk stratified screening to succeed as a cost-effective screening strategy amongst the strategies modelled, and also enables comparison with more realistic scenarios.

Scenario 2: “PRAGMATIC”: This scenario assumes the use of currently available PRSs for European ancestries (assumed to account for 90% of the modelled cohort), as well as for the two other smaller ancestries respectively accounting for 5% each of the cohort. The “true” PRS is not known in this scenario. We didn’t make any assumptions on how many SNPs would be included in this panel, and assumed that the cost of obtaining and using this PRS is £50, which is broadly similar to estimates in the Dixon et al systematic review (20).

### 2.3 Model parameters

We updated Keeney et al (29) to permit the comparison of different ways of including PRSs in PCa screening. We used data from NICE (32) on the costs of PSA testing, and otherwise relied on costs reported in Keeney et al, but inflated these costs from 2019/2020 price levels to 2022/2023 prices using the NHS Cost Inflation Index (33). Costs and utilities were discounted at 3.5% per annum. Utility values were based on the Patient-Oriented Prostate Utility Scale-Utility (PORPUS-U) instrument (34, 35). As a sensitivity analysis, we re-ran the results using EQ-5D-3L utility values.

Other aspects of the model are the same as those used and reported by Keeney et al including with respect to test accuracy, mortality rates and other clinical parameters. The parameters that distinguish each of the PRS scenarios from each other are as described above: the “pragmatic” scenario does not assume knowledge of the “true” PRS, and the PRS test itself costs £50 rather than £0.

### 2.4 Cost-effectiveness analysis

We compared all strategies to a “no screen” comparator. We calculated incremental cost-effectiveness ratios and created a representation of the cost-effectiveness frontier on which the cost-effectiveness of all alternative screening strategies was compared. Effectiveness was measured as quality adjusted life years (QALYs), combining estimates of both quality of life and mortality.

Incremental cost effectiveness ratios (ICERs) were calculated and compared to no screening for all strategies. Strategies on the frontier are considered cost-effective for different values of a cost-effectiveness threshold, whereas strategies within the frontier are dominated with respect to either or both of cost and effectiveness on the frontier. We considered all strategies against a cost-effectiveness threshold of £20,000 per QALY. The analysis was conducted and reported in accordance with the Consolidated Health Economic Evaluation Reporting Standards 2022 (CHEERS) guidance (36). A completed CHEERS checklist is provided in Supplementary Material Table S5.

### 2.5 Data and code

Data and code used to implement the model are available at: https://github.com/mclements/prostata and https://github.com/mclements/prostata/blob/develop/inst/doc/padraig.zip.

## 3 Results

The incremental lifetime cost-effectiveness of all strategies in relation to the cost-effectiveness frontier is represented in Figure 1 and in Table 2. Table 2 also reports additional information including lifetime PSA tests, biopsies, and risks of PCa diagnoses and risk of PCa deaths. As the results are from a lifetime perspective, total lifetime PCa diagnoses or PCa deaths can be multipled by the cohort size of 10 million men to get the absolute numbers of such outcomes, eg 1.57 million PCa diagnoses under no screening and 564,000 PCa deaths.

**Figure 1:**
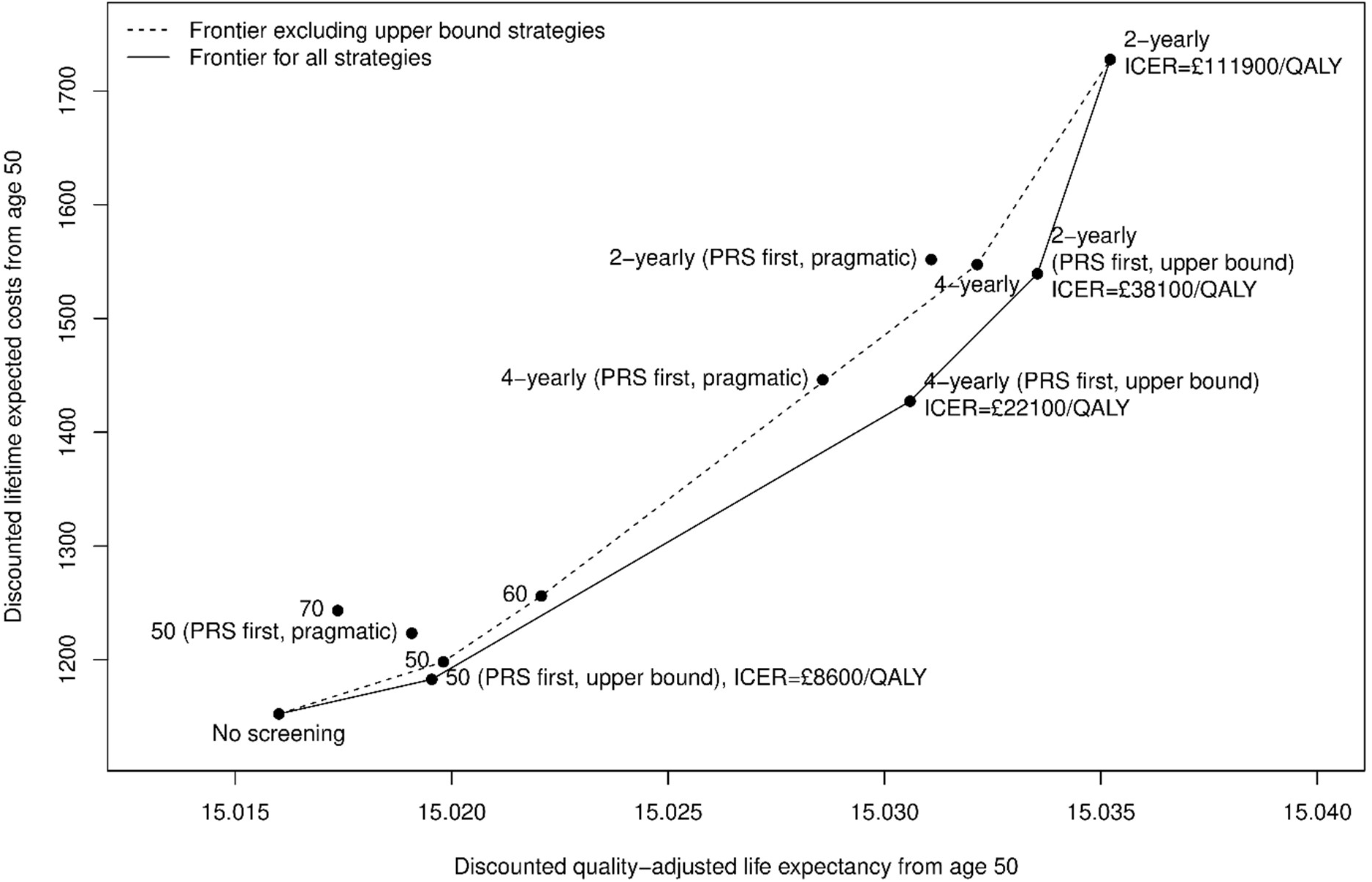
Cost-effectiveness acceptability frontier. Note to Figure 1: To enhance readability, ICERs for strategies within the efficiency frontier are not shown in the graph but are provided in Table 2 below. Table S1 in the supplementary material reports comparisons that exclude the “upper bound” PRS scenarios.

Table 1 shows that no screening has the lowest discounted cost of all strategies (£1,152), and the fewest PCa-related lifetime biopsies (0.298), but also presents the highest risk of PCa death (5.6%), and the lowest QALYs (15.016). It is important to note that the difference in QALYs between no screening and all other strategies is very narrow, indicating that any screening strategy in this cohort will likely have a small impact on this outcome.

**Table 1.** Key cost and quality-of-life model parameters.

| Resource | Unit Cost (2022/23 prices) | Lower 95% CI | Upper 95% CI | Notes |
| --- | --- | --- | --- | --- |
| PSA test | GBP 31.22 | GBP 25.29 | GBP 37.15 | From NICE (37) |
| Biopsy (systematic/MRI targeted) | GBP 653.63 | GBP 523.13 | GBP 784.13 | Updated from Keeney et al |
| Multiparametric MRI | GBP 381.38 | GBP 304.88 | GBP 457.88 | Updated from Keeney et al |
| Assessing suspected prostate cancer | GBP 613.13 | GBP 490.50 | GBP 735.75 | Updated from Keeney et al |
| Prostatectomy | GBP 11,034.00 | GBP 8,826.75 | GBP 13,241.25 | Updated from Keeney et al |
| Radiation therapy | GBP 7,269.75 | GBP 5,816.25 | GBP 8,723.25 | Updated from Keeney et al |
| Active surveillance (yearly) | GBP 649.13 | GBP 519.75 | GBP 778.50 | Updated from Keeney et al |
| Palliative care/terminal illness | GBP 8,305.88 | GBP 6,644.25 | GBP 9,967.50 | Updated from Keeney et al. Terminal care assumed to last for six months; palliative care for the 6-30 months prior. |
| Health states | Utility value | Lower 95% CI | Upper 95% CI | Duration of state |
| PSA test | 0.99 | 0.99 | 1 | 1 week |
| Biopsy (SBx, TBx, TBx/SBx) | 0.9 | 0.87 | 0.94 | 3 weeks |
| Cancer diagnosis | 0.8 | 0.75 | 0.85 | 1 month |
| Active surveillance | 0.9 |  |  | 7 year |
| Radical prostatectomy (part 1) | 0.86 | 0.76 | 0.96 | 2 months |
| Radical prostatectomy (part 2) | 0.90 | 0.84 | 0.97 | 10 months |
| Radiation therapy (part 1) | 0.89 | 0.87 | 0.91 | 2 months |
| Radiation therapy (part 2) | 0.92 | 0.90 | 0.94 | 10 months |
| Androgen deprivation therapy and chemotherapy | 0.80 | NR | NR | 18 months |
| Post recovery period | 0.93 | 0.91 | 0.95 | 9 years |
| Palliative therapy | 0.68 | 0.64 | 0.71 | 12 months |
| Terminal illness | 0.4 | NR | NR | 6 months |

Note to Table 1 NR: Not reported..

Under anti-conservative “upper bound” scenarios that assume knowledge of an unobtainable “true” PRS, fewer PSA tests and biopsies are required to achieve a slight reduction in PCa mortality compared to no screening. For instance, the 4-yearly PRS-first (upper bound) strategy involves around 2 PSA tests and 0.33 biopsies per person, with a PCa death risk of 5.02%, compared to 5.64% with no screening. The more realistic “pragmatic” PRS strategies involve slightly more testing and have a smaller impact on PCa mortality reduction than “upper bound” strategies. For example, the 4-yearly PRS- first (pragmatic) strategy results in a similar number of PSA tests (2.04) and biopsies (0.33), but the mortality reduction is more modest at 5.11%. The cost-effectiveness of each strategy is summarised in the ICERs reported in Table 2 and in Figure 1.

**Table 2.**
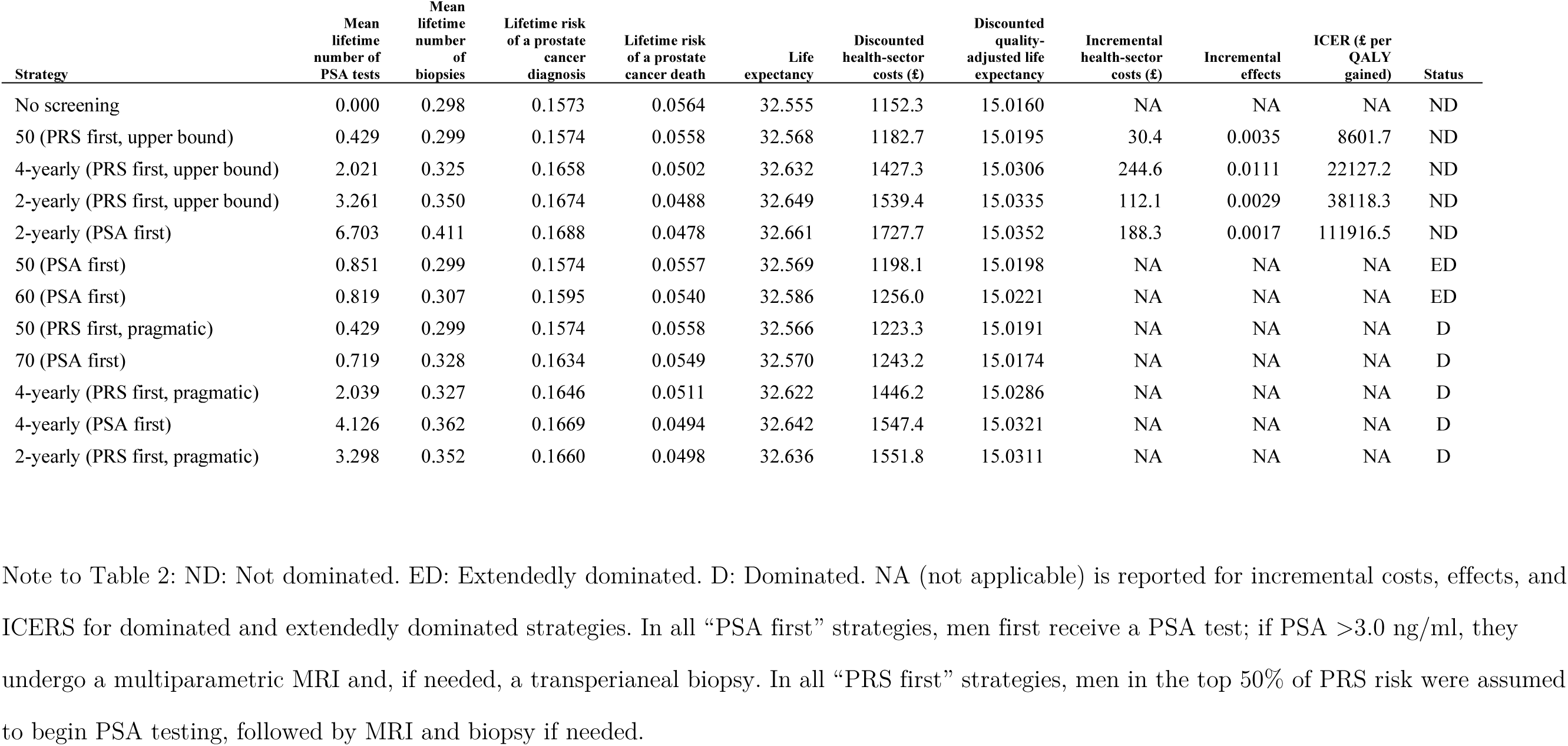
Lifetime expected outcomes and cost-effectiveness from age 50 years.

In relation to the cost-effectiveness frontier (Figure 1), both no screening and a single one off-screen at age 50 are on the cost-effectiveness frontier, and fall beneath a £20,000 per QALY cost-effectiveness threshold.

The indicates that a cost-free “ideal” PRS from the “upper bound” scenario could be cost-effective, while a more realistic PRS from the “pragmatic scenario”, may not offer greater cost-effectiveness compared to a single screen (using PSA) at age 50 or even compared to no screening at all. The findings also suggest that improved PRS stratification would improve cost-effectiveness, as evidenced from a hypothetical movement from the “pragmatic” PRS screening strategy to the unobtainable “upper bound” strategy on the cost-effectiveness frontier. However, this alone does not imply that investing in such improvements would be the most efficient use of resources, as PRSs may never sufficiently approach or reach the performance of the “ideal” or true PRS included in the “upper bound” scenarios.

Results excluding the less realistic “upper bound” scenario are presented in Figure S1 (the frontier) and Table S1 in supplementary material. Results were broadly similar when using EQ-5D instead of PORPUS-U values for quality-of-life parameters, although ICERS increased more sharply compared to no screening when using EQ-5D utility inputs. As before, no screening was on the cost-effectiveness frontier (Figure S1 in supplementary material), and only PRS-first scenarios in the “upper bound” scenario were likely to be cost-effective, and all of the more realistic PRS strategies from the “pragmatic” scenario were unlikely to be cost-effective. Tables S2 and S3 report results from using EQ-5D rather than PORPUS-U for utility values.

## 4 Discussion

This modelling study found that incorporating realistic PRSs into PCa screening is not cost-effective. This partly reflects the very low cost of PSA testing compared to polygenic risk stratification as a first or early step in triggering further investigations in the diagnostic workup. While an affordable, highly predictive, and widely available polygenic risk score may ultimately supplement some of these tests, it is not clear from this modelling evidence that this is imminent, nor that they would displace existing tests.

### 4.1 Related literature

Amongst the screening modalities we studied were “PRS first” strategies. A recent study (37) also adopted a PRS first strategy. In this UK-based study, men aged 55 to 69 were recruited from primary care, and those in the top 10% of a 130-SNP PRS for incident PCa were invited to undergo MRI and biopsy, regardless of PSA levels. The PRS-first approach resulted in more overdiagnosis than PSA testing alone or MRI alone. The study did not have a control arm, was limited in follow-up, and didn’t report on PCa mortality, didn’t compare these strategies to other plausible uses of PRSs in screening or to other screening modalities amongst the recruited men, and economic outcomes including cost and quality-of-life were not reported.

The TRANSFORM trial (38) was announced in the UK in 2024 and is due to recruit its first participants in 2025. In the first stage of this study, approximately 12,500 men living in the UK will be randomized to screening modalities including MRI scans, PRS stratification and PSA blood tests. In the second stage of the study, the most promising approaches identified in the first stage will be studied in up to 300,000 participants. The evidence from our modelling suggests that the case albeit the case for “PRS-first” study arms in TRANSFORM, if these are to be considered, may not be compelling

There are a small number of other PCa screening studies undertaken from an NHS perspective that account for polygenic risk. The present study is based on a revised and updated version of the Keeney et al model, in which the two polygenic risk stratified screening strategies involved screening every two four years starting at the age when a person’s 10-year risk of developing PCa reaches 7.5%,. Neither of these strategies was on the cost-effectiveness frontier, being less cost-effective than a one-off screen or no screening, but were potentially more cost-effective than simple age-based screening strategies.

Callender et al (39) concluded that polygenic risk stratification could reduce overdiagnosis and have comparable mortality benefits to age-based screening, and concluded that the optimal threshold for this form of risk-stratified screening depends on societal preferences regarding the trade-off between screening benefits and harms. Callender et al (40) adapted this model in an evaluation of cost-effectiveness associated with MRI before biopsy compared with biopsy-first screening. This study concluded that improvements in cost-effectiveness under a MRI-first diagnostic pathway were more significant with risk-stratified screening based on age and polygenic risk.

Huntley et al (41) considered cancer detection rates for five PRS quintiles, and modelled the introduction of PCa screening for men aged 60-69. Of these men, 46% of annual PCa cases would arise in the highest risk PRS quintile. The study estimates that extending screening to the PRS-defined high-risk quintile of men aged 60–69 could avert up to 158 deaths annually, compared to 95 annual deaths averted under unstratified screening. The cost-effectiveness of different strategies were not studied, and modelled benefits were anti-conservative in that uptake of PRS testing will be incomplete, the presence of interval cancers was not modelled, and given reduced PRS applicability in non-European populations. The costs and cost-effectiveness of PRS-stratified screening were not studied.

### 4.2 Strengths

We used an established PCa screening microsimulation to model potential impacts of PRS-informed risk stratification in three scenarios. This model used a detailed natural history model, and is well calibrated (29) to mortality estimates from external sources such as the ERSPC and CAP trials. As a validation of the natural history model, we simulated for a reconstruction of the CAP trial to 15 years. We estimated a mortality rate ratio of 0.94 for the screening arm compared with the control arm. This point estimate is consistent with the estimated mortality rate ratio from Martin et al (5), who estimated a mortality rate ratio of 0.92 (95% CI: 0.85, 0.99)

We considered different possible implementations of polygenic risk scores. We compared a “pragmatic” scenario to an optimistic “upper bound” scenario. This approach permitted inferences about potential upper bounds of gains from screening using these scores. These scores attempted to account for differential ancestry and uncertainty over the costs of testing, two significant issues identified in a systematic review (20) regarding the use of polygenic risk scores in cancer screening.

### 4.3 Limitations

This work has a number of limitations. We attempted to account for some of the more consequential features of polygenic risk stratification but had to make several assumptions concerning the potential scale of some variables influencing screening cost-effectiveness.

These included the costs of PRSs, an assumption that they would be widely available and acceptable to men participating in screening, and assumptions concerning their accuracy in general and in relation to different ancestral backgrounds. In the “pragmatic” scenario, we based PRS costs on estimates from studies included in the Dixon et al. (20) systematic review; however, real-world costs may be higher, especially if downstream effects—such as increased primary care visits for reassurance—add costs beyond the screening programme.

Ancestry enters the model through the influence of ancestry-specific polygenic risk on frailty. While we included different ancestries in the model, this was limited to just three different ethnicities. One of these ancestries has higher relative risk (compared to the base European ancestry) to reflect elevated risk amongst black men. The other has lower relative risk to reflect associations of incident disease with (very broadly defined) Asian ancestry. In reality, ancestral backgrounds cannot be neatly categorized into these three broad groups, but this was done for pragmatic modelling purposes. In the absence of evidence on the modelled ethnic groups, we did not assume or model any ancestral differences in test performance, disease progression, or mortality outcomes. We also modelled the PRS as affecting only the risk of incident PCa, with no impact on disease progression. Our PRSs are hypothetical and do not reflect individual-level data.

We also considered but did not model other potential impacts of PRS on cost-effectiveness parameters. In particular, there is scant evidence to suggest PRSs increase or decrease responses to screening invitations e.g. (17). There is also little evidence knowledge of own PRS is associated with changes of quality of life, or rates of consultation in primary care, although such knowledge may be associated with transitory impacts on anxiety for some individuals (42).

Our different screening strategies did not attempt to identify an “optimal” risk threshold for screening that accounts for patient preferences concerning trade-offs between overdiagnosis, quality of life, and death. Modelled screening strategies attempted to reflect realistic practical implementations of PCa screening, and necessarily modelled only a limited number of ways to include PRS-based screening.

We assumed that PSA levels at or above 3ng/ml would trigger investigation. At the time of writing, this differs from NICE guidelines (43) for the use of PSA in the assessment of suspected PCa, which are age dependent. For men under 40, guidelines recommend relying on clinical judgment, while levels above 2.5 µg/L for ages 40–49, 3.5 µg/L for 50– 59, 4.5 µg/L for 60–69, and 6.5 µg/L for 70–79 merit further investigation.

#### Uncertainty and probabilistic sensitivity analysis

We do not report probabilistic sensitivity analysis. While the use of common random numbers does reduce variance in the model, and ensures comparability in the simulated experiences of a very large cohort (10 million individuals), it is still possible that parameter uncertainty may have a bearing on the interpretation of results from our model. However, the core parameters that materially differ between strategies (other than those that define the strategies themselves such as screening intervals) are the costs of PRSs and the accuracy of these scores. These features are both accounted for in our “upper bound” and “pragmatic” scenarios, which could be interpreted as a type of deterministic sensitivity analysis in which both of these key model parameters are varied.

Probabilistic sensitivity analysis also does not address structural uncertainty, such as the appropriate thresholds at which certain model inputs like PSA or PRS scores would trigger further investigation or alter screening pathways. These are questions of model design and structure rather than parameter uncertainty. While some structural uncertainty is captured indirectly through the inclusion of multiple screening strategies, it is not feasible to model every possible combination of thresholds or pathways. As a result, only a limited number of plausible strategies can be included.

In addition, the computational burden of conducting even a modest number of PSA runs (e.g., 1,000 iterations) on a cohort of 10 million individuals is substantial. Despite potential efficiency gains through techniques like meta-modelling (e.g., reducing the size of the simulated cohort or using surrogate models), the overall resource demands remain considerable.

### 4.4 Conclusions

Our objective in this paper was to characterise the potential impact of PRSs on the cost-effectiveness of stratified PCa screening. Realistic implementations of PRS-based risk stratification are unlikely to achieve cost-effectiveness at present.

## Declarations

The views expressed are those of the authors and not necessarily those of Cancer Research UK.

## Funding

This work was supported by Cancer Research UK under grant number C18281/A29019. PD and RMM received support from a Cancer Research UK (C18281/A29019) programme grant (the Integrative Cancer Epidemiology Programme). MC received funding from the Swedish Research Council (Vetenskapsrådet; VR 2022- 00684 and the Swedish eScience Research Centre), Swedish Cancer Society (Cancerfonden; CAN21/1512); Swedish Prostate Cancer Foundation (Prostatacancerförbundet); Region Stockholm (ALF grant FoUI-987651); and the European Commission (HEAP grant No. 874662).

RMM is a National Institute for Health Research Senior Investigator (NIHR202411). RMM is also supported by the NIHR Bristol Biomedical Research Centre which is funded by the NIHR (BRC-1215-20011) and is a partnership between University Hospitals Bristol and Weston NHS Foundation Trust and the University of Bristol. Department of Health and Social Care disclaimer: The views expressed are those of the author(s) and not necessarily those of the NHS, the NIHR or the Department of Health and Social Care.

The CAP trial was funded by grants C11043/A4286, C18281/A8145, C18281/A11326, C18281/A15064; and C18281/A24432 from Cancer Research UK. The UK Department of Health, National Institute of Health Research provided partial funding.

The ProtecT trial was funded by project grants 96/20/06 and 96/20/99 from the UK National Institute for Health Research, Health Technology Assessment Programme.

## Conflict of interest statement

The authors declare no conflicts of interest. RM has received other funding from Cancer Research UK to evaluate the long-term effectiveness and cost-effectiveness of population-based screening and treatment for prostate cancer: the CAP and ProtecT randomized controlled trials. JA has received honoraria from Accord, Astellas, AstraZeneca, Bayer, BXTA, Janssen (J&J Innovative Medicine), Merck.

## Supplementary Material

**Supplementary Table S1:**
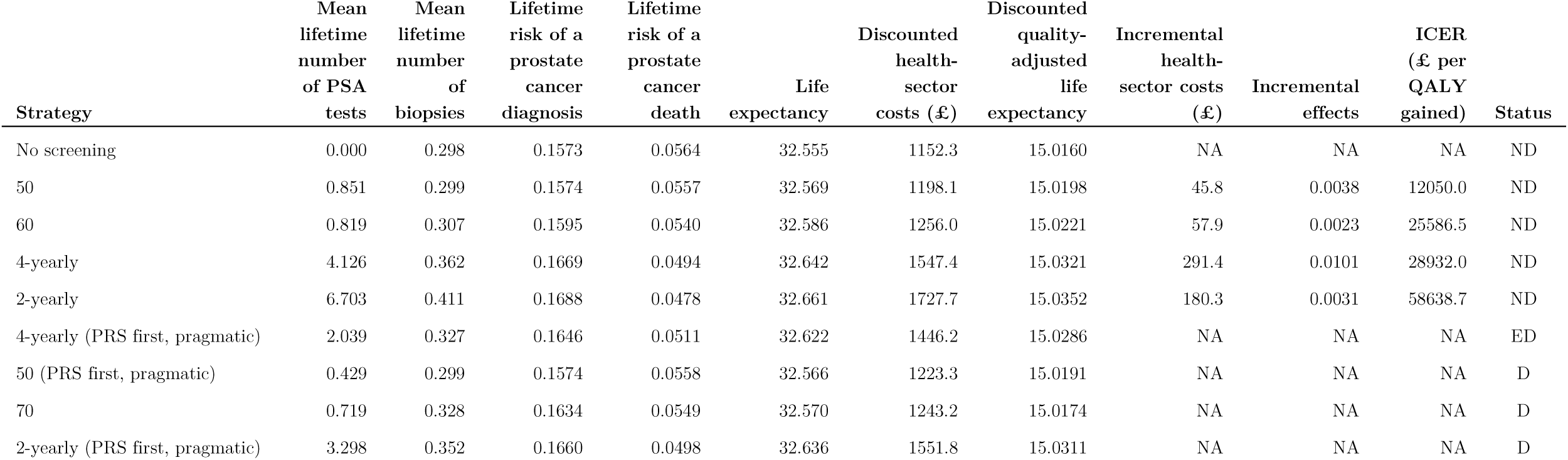
Lifetime expected outcomes and cost-effectiveness from age 50 excluding the upper bound strategies.

**Supplementary Table S2:**
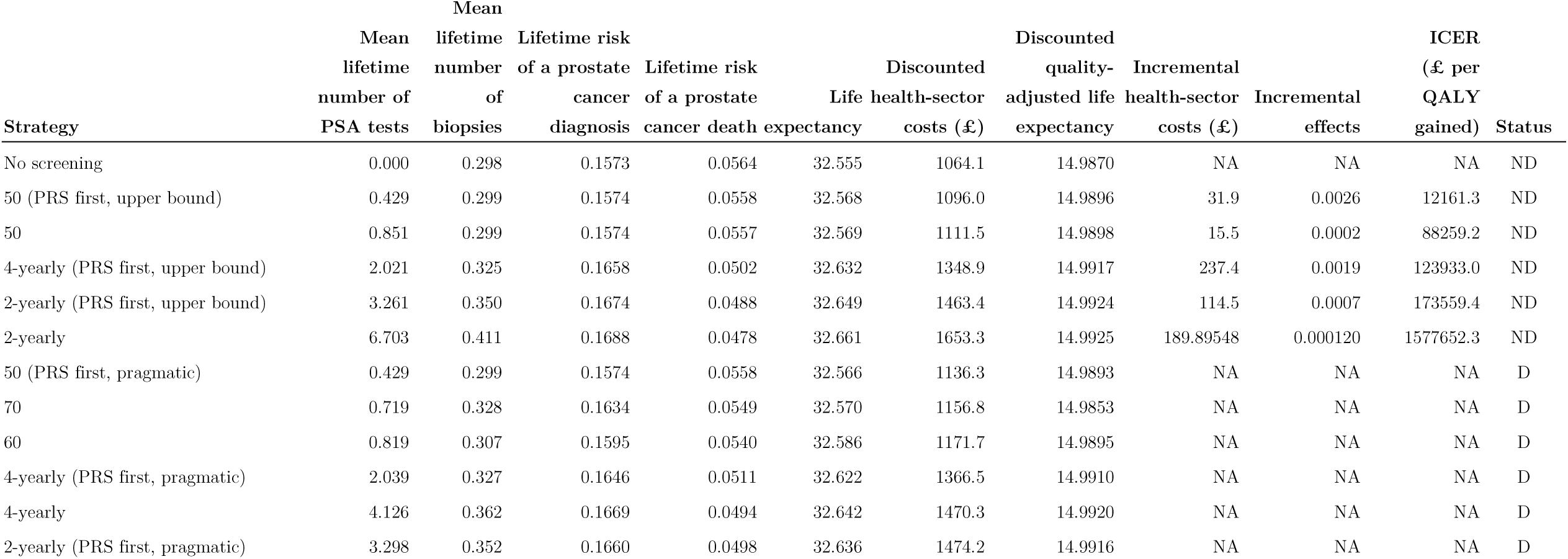
Lifetime expected outcomes and cost-effectiveness from age 50, using EQ-5D rather than PORPUS-U for utility values.

**Supplementary Table S3:**
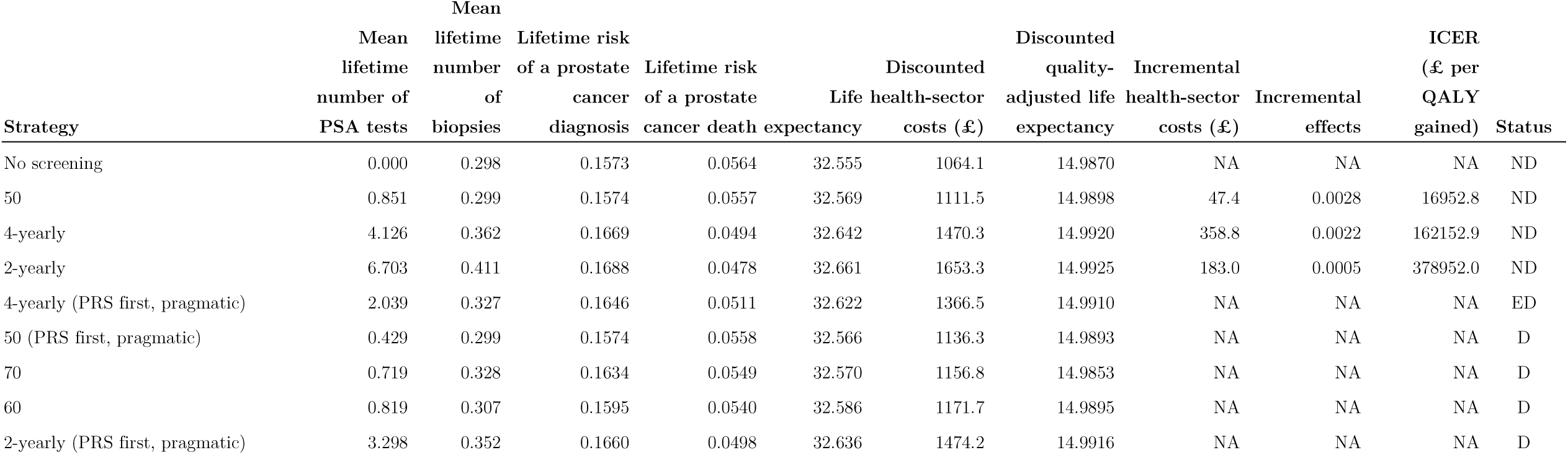
Lifetime expected outcomes and cost-effectiveness excluding the upper bound strategies from age 50, using EQ-5D rather than PORPUS-U for utility values.

**Supplementary Table S4:** EQ-5D health state values used as alternative to PORPUS-U values.

| Health states | Utility value (EQ-5D) | Lower 95% CI | Upper 95% CI | Duration of state |
| --- | --- | --- | --- | --- |
| PSA test | 0.99 | 0.99 | 1 | 1 week |
| Biopsy (SBx, TBx, TBx/SBx) | 0.9 | 0.87 | 0.94 | 3 weeks |
| Cancer diagnosis | 0.8 | 0.75 | 0.85 | 1 month |
| Active surveillance | 0.90 | 0.85 | 0.95 | 7 year |
| Radical prostatectomy (part 1) | 0.83 | 0.73 | 0.91 | 2 months |
| Radical prostatectomy (part 2) | 0.89 | 0.88 | 0.91 | 10 months |
| Radiation therapy (part 1) | 0.82 | 0.75 | 0.88 | 2 months |
| Radiation therapy (part 2) | 0.83 | 0.81 | 0.85 | 10 months |
| Androgen deprivation therapy and chemotherapy | 0.73 | 0.68 | 0.77 | 18 months |
| Post recovery period | 0.86 | 0.84 | 0.88 | 9 year |
| Palliative therapy | 0.62 | 0.58 | 0.66 | 12 months |
| Terminal illness | 0.4 | NR | NR | 6 months |

**Supplementary Table S5.** CHEERS Checklist.

| Section/topic | Item No | Guidance for reporting | Reported in section |
| --- | --- | --- | --- |
| **Title** |  |  |  |
| Title | 1 | Identify the study as an economic evaluation and specify the interventions being compared. | P1 |
| **Abstract** |  |  |  |
| Abstract | 2 | Provide a structured summary that highlights context, key methods, results, and alternative analyses. | P2 |
| **Introduction** |  |  |  |
| Background and objectives | 3 | Give the context for the study, the study question, and its practical relevance for decision making in policy or practice. | P3-4 |
| **Methods** |  |  |  |
| Health economic analysis plan | 4 | Indicate whether a health economic analysis plan was developed and where available. | P4 |
| Study population | 5 | Describe characteristics of the study population (such as age range, demographics, socioeconomic, or clinical characteristics). | P4-6 |
| Setting and location | 6 | Provide relevant contextual information that may influence findings. | P4-6 |
| Comparators | 7 | Describe the interventions or strategies being compared and why chosen. | P6-7 |
| Perspective | 8 | State the perspective(s) adopted by the study and why chosen. | P6 |
| Time horizon | 9 | State the time horizon for the study and why appropriate. | P4 |
| Discount rate | 10 | Report the discount rate(s) and reason chosen. | P7 |
| Selection of outcomes | 11 | Describe what outcomes were used as the measure(s) of benefit(s) and harm(s). | P7-8 |
| Measurement of outcomes | 12 | Describe how outcomes used to capture benefit(s) and harm(s) were measured. | P7-8 |
| Valuation of outcomes | 13 | Describe the population and methods used to measure and value outcomes. | P7-8 |
| Measurement and valuation of resources and costs | 14 | Describe how costs were valued. | P7-8 |
| Currency, price date, and conversion | 15 | Report the dates of the estimated resource quantities and unit costs, plus the currency and year of conversion. | P7-8 |
| Rationale and description of model | 16 | If modelling is used, describe in detail and why used. Report if the model is publicly available and where it can be accessed. | P4-10 |
| Analytics and assumptions | 17 | Describe any methods for analysing or statistically transforming data, any extrapolation methods, and approaches for validating any model used. | P4-10 |
| Characterising heterogeneity | 18 | Describe any methods used for estimating how the results of the study vary for subgroups. | NA |
| Characterising distributional effects | 19 | Describe how impacts are distributed across different individuals or adjustments made to reflect priority populations. | NA |
| Characterising uncertainty | 20 | Describe methods to characterise any sources of uncertainty in the analysis. | P4-10, P19-20 |
| Approach to engagement with patients and others affected by the study | 21 | Describe any approaches to engage patients or service recipients, the general public, communities, or stakeholders (such as clinicians or payers) in the design of the study. | NA |
| Study parameters | 22 | Report all analytic inputs (such as values, ranges, references) including uncertainty or distributional assumptions. | P8-9 |
| Summary of main results | 23 | Report the mean values for the main categories of costs and outcomes of interest and summarise them in the most appropriate overall measure. | P10-14 |
| Effect of uncertainty | 24 | Describe how uncertainty about analytic judgments, inputs, or projections affect findings. Report the effect of choice of discount rate and time horizon, if applicable. | P5, P19-20 |
| Effect of engagement with patients and others affected by the study | 25 | Report on any difference patient/service recipient, general public, community, or stakeholder involvement made to the approach or findings of the study | NA |
| Study findings, limitations, generalisability, and current knowledge | 26 | Report key findings, limitations, ethical or equity considerations not captured, and how these could affect patients, policy, or practice. | P14-20 |
| **Other relevant information** |  |  |  |
| Source of funding | 27 | Describe how the study was funded and any role of the funder in the identification, design, conduct, and reporting of the analysis | P21 |
| Conflicts of interest | 28 | Report authors conflicts of interest according to journal or International Committee of Medical Journal Editors requirements. | P21 |

**Figure S1:**
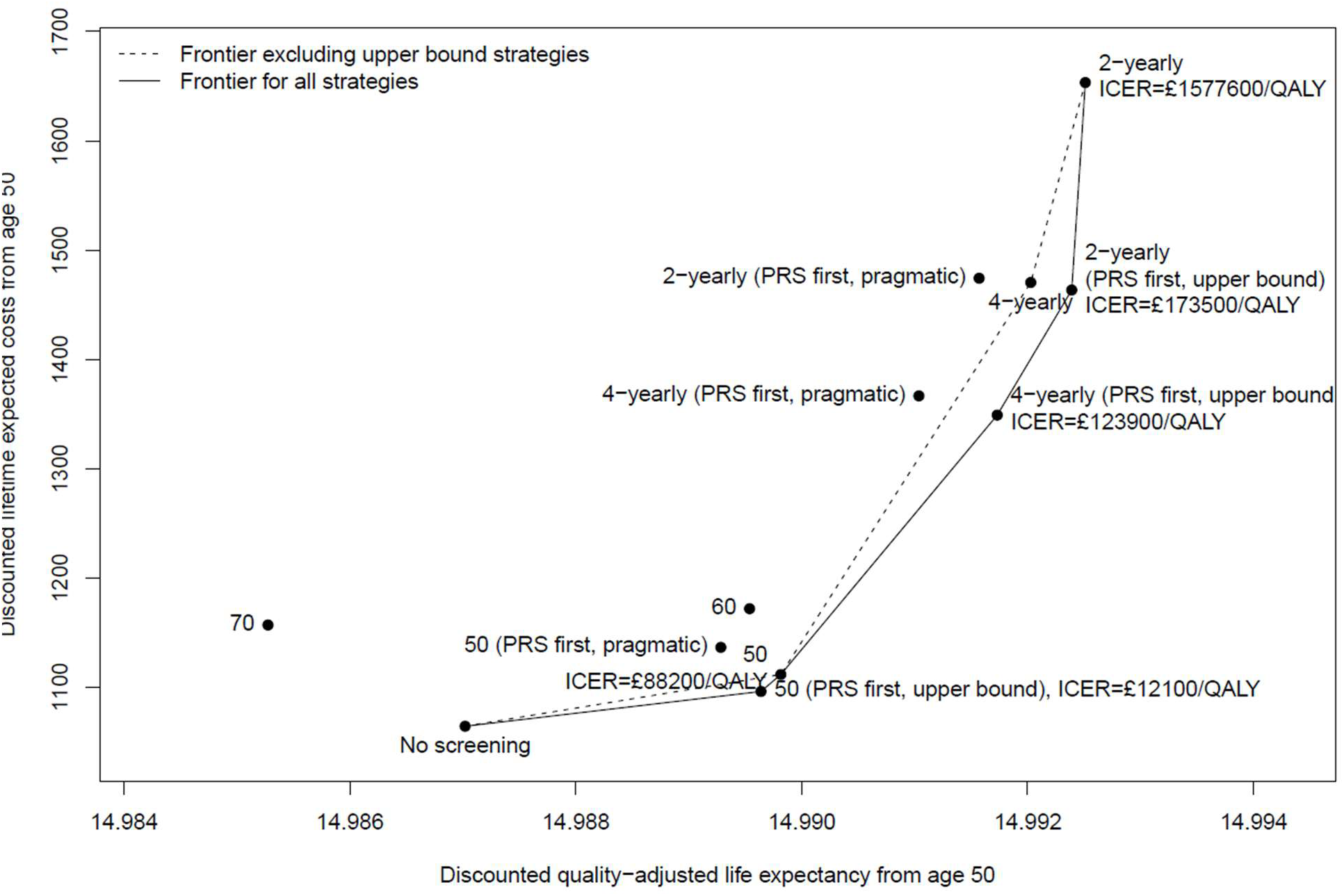
Cost-effectiveness acceptability frontier using EQ-5D utility values.

